# Do Amyloid Trajectories Reach a Physiologic Ceiling? Evidence from Iterative Approximation and Simulation

**DOI:** 10.64898/2026.04.14.26350359

**Authors:** Jason R. Gantenberg, Renaud La Joie, Margo B. Heston, Sarah F. Ackley, the Alzheimer’s Disease Neuroimaging Initiative

**Affiliations:** Department of Health Services, Policy, and Practice, Brown University, 121 S. Main St., Providence, RI 02903; Department of Epidemiology, Brown University, 121 S. Main St., Providence, RI 02903; Edward and Pearl Fein Memory and Aging Center, Weill Institute for Neurosciences, Department of Neurology, University of California San Francisco, 1651 4th St Suite 212, San Francisco, CA 94158

## Abstract

Qualitative models of Alzheimer’s pathology often posit that amyloid accumulation follows a sigmoid curve, indicating that the rate of deposition wanes over time. Longitudinal PET data now allow us to investigate amyloid accumulation trajectories with greater detail and over longer follow-up periods. We combine inferences from simulated amyloid trajectories, empirical PET data from the Alzheimer’s Disease Neuroimaging Initiative (ADNI), and the sampled iterative local approximation algorithm (SILA) to assess whether amyloid accumulation reaches a physiologic ceiling. We find that SILA reliably detects a ceiling, when present, across a range of simulated scenarios that impose a sigmoid shape. When fit to empirical data from ADNI, however, SILA does not appear to indicate the presence of a ceiling. Thus, we conclude that amyloid trajectories may not reach a physiologic ceiling during the stages of Alzheimer’s disease typically observed while patients remain under follow-up in cohort studies. Fits using SILA indicate that illustrative models of biomarker cascades, while useful tools for conceptualizing and interrogating pathologic processes, may not represent the shapes of amyloid trajectories accurately.

**Summary for General Public:** Amyloid, a protein implicated in Alzheimer’s disease, is thought to reach a plateau in the brain, but methods that estimate how amyloid changes over time suggest it grows unabated. Gantenberg et al. use one such method and simulations to argue that amyloid does not reach a plateau during the typical course of Alzheimer’s.

## Introduction

Research has been increasingly focused on modeling longitudinal biomarker trajectories to monitor the pathophysiology of neurodegenerative diseases.^1–7^

When phenomenological models positing sigmoid biomarker progression in AD emerged around the early 2010s, evidence for a physiologic ceiling in amyloid accumulation was largely indirect, as amyloid-PET was still relatively new.^8,9^ An increase followed by a decrease in the rate of amyloid accumulation produces a sigmoid curve,^10^ and prior research, usually in small samples, sometimes appears to corroborate this pattern.^5,10^ A neuropathological study suggested an amyloid ceiling, finding that amyloid burden was not associated with disease duration in AD cases^11^; however, autopsy studies involve selection biases that prohibit applying inferences to living individuals.^12^ Another found that a sigmoid fit better than a line when regressing cognition on CSF amyloid-beta.^13^ Historically, longitudinal amyloid measurements have been available over short follow-up periods^5,10^ relative to the decades-long course of AD.

More recent prospective studies have collected repeated amyloid-PET measures over significantly longer time frames, enabling more robust inferences about amyloid accumulation trajectories. Publications based on these studies continue to report mixed findings regarding the existence of a pathophysiological amyloid ceiling.^1,4,5,14^ The sampled iterative local approximation (SILA) algorithm is a recent and popular method that has been used to model both PET and blood biomarker trajectories in Alzheimer’s disease (AD).^1,15,16^ Briefly, SILA produces a mean trajectory that can be used to estimate the age of amyloid positivity onset (AAPO) for a given individual in a sample.^1^ Its outputs can also be used as predictors or outcomes in subsequent modeling.^15,17^ By aligning measurements across individuals sampled at different points along their amyloid trajectory, SILA may allow us to accurately infer the average accumulation trajectory pre-disease onset and through the course of AD. However, SILA has not been systematically stress-tested to determine whether it reliably recovers the qualitative shape of underlying amyloid trajectories or captures potential inter-individual heterogeneity in accumulation rates.

With data from the Alzheimer’s Disease Neuroimaging Initiative (ADNI), we simulated hypothetical amyloid trajectories using realistically spaced amyloid PET measurements. To demonstrate that SILA reliably detects the qualitative shape of the population average amyloid trajectory across a range of scenarios, we varied the generative rules that governed the shape, rate, and inter-individual heterogeneity of these curves. We focus on SILA’s ability to detect a physiologic ceiling under data constraints similar to those that exist in ADNI.

## Methods

### ADNI Cohort

Data used in the preparation of this article were obtained from the Alzheimer’s Disease Neuroimaging Initiative (ADNI) database (adni.loni.usc.edu). The ADNI was launched in 2003 as a public-private partnership, led by Principal Investigator Michael W. Weiner, MD. The primary goal of ADNI has been to test whether serial magnetic resonance imaging (MRI), positron emission tomography (PET), other biological markers, and clinical and neuropsychological assessment can be combined to measure the progression of mild cognitive impairment (MCI) and early Alzheimer’s disease (AD). For up-to-date information, see www.adni-info.org. Imaging and participant data were downloaded February 13, 2026 for the present analysis.

PET imaging procedures and processing are described in detail elsewhere.^18^ Amyloid burden was quantified using ^18^F-florbetapir, ^18^F-florbetaben, or ^18^F-flutafuranol PET by the ADNI PET core at the University of California, Berkeley. Centiloid values used in the current manuscript are derived from an MRI-based processing pipeline of the PET scans (6 mm effective resolution). We retrieved information on participants’ ages and cognitive diagnoses from the PTDEMOG and DXSUM datasets, respectively.

Information regarding patient subsets used in our estimation procedures appears in the following sections and in **Supplementary Table 1**.

### Empirical SILA Fit

SILA is an algorithm developed by Betthauser et al.^1^ that iteratively samples biomarker observations and applies local regressions to infer a population average trajectory based on partially observed individual-level trajectories. We used the *silaR* package (https://github.com/Betthauser-Neuro-Lab/silaR) to fit SILA on empirical PET data from ADNI participants with at least 2 Centiloid measurements (3,539 amyloid-PET in 1,141 participants). From this fit we extracted SILA’s predictions for AAPO and amyloid trajectories relative to AAPO. Characteristics of the empirical sample used to fit SILA appear in **Table 1**.

**Table 1.**
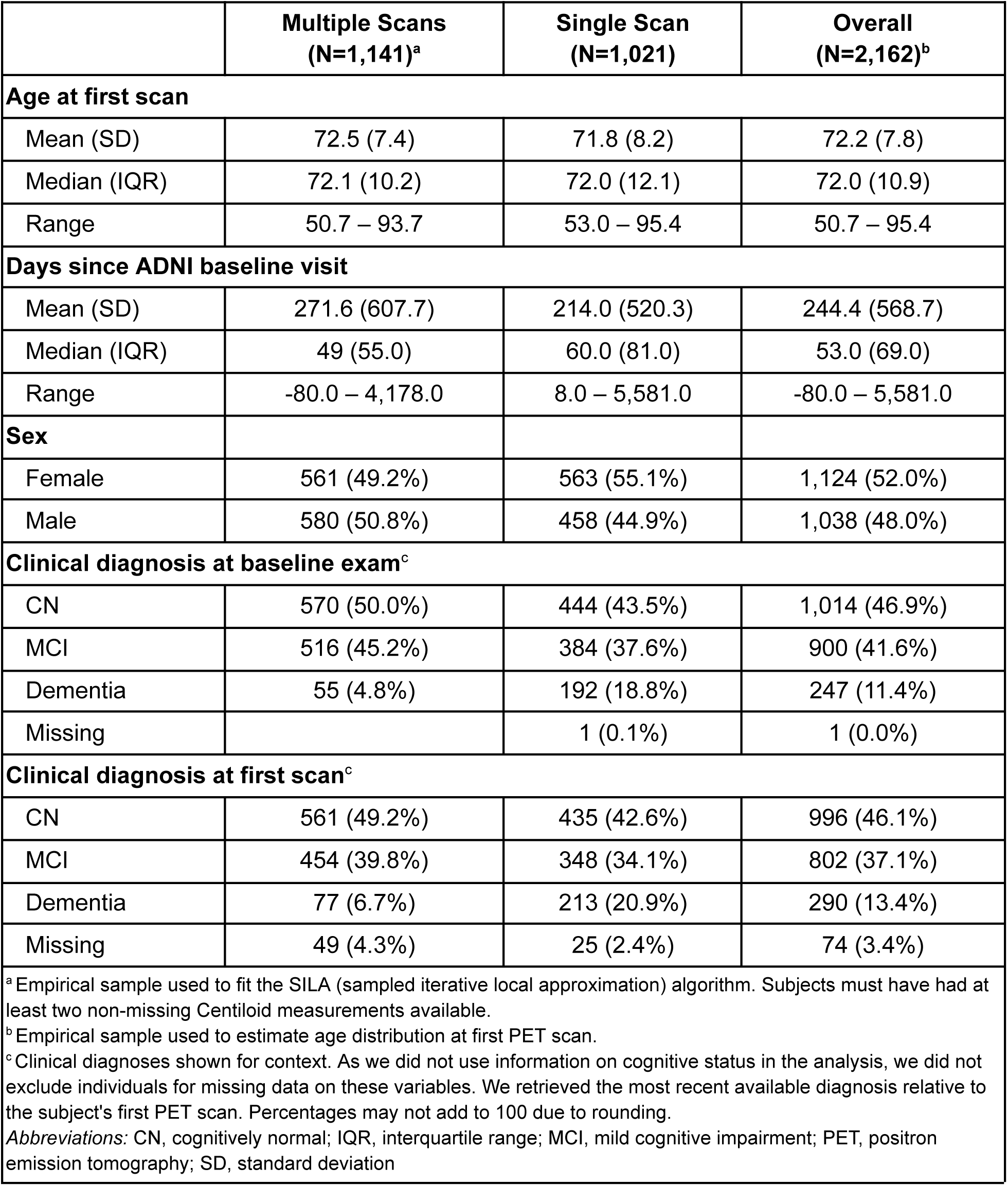
Characteristics of ADNI participants at their first PET scan.

|  | **Multiple Scans**<br>**(N=1,141)<sup>a</sup>** | **Single Scan**<br>**(N=1,021)** | **Overall**<br>**(N=2,162)<sup>b</sup>** |
| --- | --- | --- | --- |
| **Age at first scan** |  |  |  |
| Mean (SD) | 72.5 (7.4) | 71.8 (8.2) | 72.2 (7.8) |
| Median (IQR) | 72.1 (10.2) | 72.0 (12.1) | 72.0 (10.9) |
| Range | 50.7 – 93.7 | 53.0 – 95.4 | 50.7 – 95.4 |
| **Days since ADNI baseline visit** |  |  |  |
| Mean (SD) | 271.6 (607.7) | 214.0 (520.3) | 244.4 (568.7) |
| Median (IQR) | 49 (55.0) | 60.0 (81.0) | 53.0 (69.0) |
| Range | -80.0 – 4,178.0 | 8.0 – 5,581.0 | -80.0 – 5,581.0 |
| **Sex** |  |  |  |
| Female | 561 (49.2%) | 563 (55.1%) | 1,124 (52.0%) |
| Male | 580 (50.8%) | 458 (44.9%) | 1,038 (48.0%) |
| **Clinical diagnosis at baseline exam<sup>c</sup>** |  |  |  |
| CN | 570 (50.0%) | 444 (43.5%) | 1,014 (46.9%) |
| MCI | 516 (45.2%) | 384 (37.6%) | 900 (41.6%) |
| Dementia | 55 (4.8%) | 192 (18.8%) | 247 (11.4%) |
| Missing |  | 1 (0.1%) | 1 (0.0%) |
| **Clinical diagnosis at first scan<sup>c</sup>** |  |  |  |
| CN | 561 (49.2%) | 435 (42.6%) | 996 (46.1%) |
| MCI | 454 (39.8%) | 348 (34.1%) | 802 (37.1%) |
| Dementia | 77 (6.7%) | 213 (20.9%) | 290 (13.4%) |
| Missing | 49 (4.3%) | 25 (2.4%) | 74 (3.4%) |
| <sup>a</sup> Empirical sample used to fit the SILA (sampled iterative local approximation) algorithm. Subjects must have had at least two non-missing Centiloid measurements available.<br><sup>b</sup> Empirical sample used to estimate age distribution at first PET scan.<br><sup>c</sup> Clinical diagnoses shown for context. As we did not use information on cognitive status in the analysis, we did not exclude individuals for missing data on these variables. We retrieved the most recent available diagnosis relative to the subject's first PET scan. Percentages may not add to 100 due to rounding.<br>*Abbreviations:* CN, cognitively normal; IQR, interquartile range; MCI, mild cognitive impairment; PET, positron emission tomography; SD, standard deviation |  |  |  |

### Simulations

We simulated 100 amyloid datasets per scenario, each for a group of 2,000 hypothetical individuals with realistic numbers and spacing of PET scans, parameterized based on ADNI. We considered two different primary shapes for the “true” amyloid trajectories: exponential and logistic. The exponential produces trajectories that grow without bound, while the logistic trajectory produces a sigmoid with fast early growth that decelerates as it hits a ceiling.^8,10^ Second, we simulated scenarios in which individuals were homogeneous or heterogeneous with respect to amyloid accumulation. Because SILA implicitly assumes inter-individual homogeneity in biomarker accumulation rate, we wished to evaluate its ability to detect a ceiling when this assumption was violated. **Supplementary Table 2** outlines the simulation parameters and accumulation trajectories we employed.

### Generating Realistic Neuroimaging Cohorts

We used ADNI data to characterize the age at which individuals receive their first amyloid-PET scan, how many scans they receive, and the timing between scans. These empirically derived patterns were then used to generate simulated individuals emulating observed scanning histories and estimated AAPOs. Specifically, we constrained simulated datasets to resemble the subset of ADNI participants with at least one PET scan in the distributions of expected number of scans, age at first PET scan, and SILA-estimated AAPO, as well as the subset of participants with ≥ 2 scans in the distribution of the number of days elapsed between scans.

We calculated the *expected number of scans* by calculating the proportions of subjects with *n* scans and used these proportions as weights to determine how many scans a simulated individual received. We calculated the mean and standard deviation of *age at first scan* and used these values to simulate from a Normal distribution. To simulate the *days elapsed between scans*, we estimated the distribution of differences between subsequent scan dates using the Sheather-Jones bandwidth and then simulated from this density. To calculate *AAPO* we extracted each subject’s SILA-estimated AAPO from the empirical sample. We fit AAPO to a gamma distribution and used this fit to generate simulated data. More information regarding the data used to estimate these parameters appears in **Supplementary Table 1**.

### Amyloid Accumulation Trajectories

*Homogeneity* in the following description refers to the assumption that all individuals in a population follow the same amyloid accumulation trajectory relative to their AAPO.

Simulated Centiloid measurements under the exponential accumulation scenarios followed

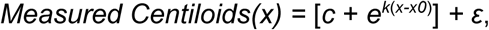

where *c* is a constant vertical offset, *k* is the rate parameter, *x* is the number of years relative to the simulated AAPO, *x0* is the calculated center of the function, and *ε* is normally distributed noise to simulate a small amount of measurement error based on empirical estimates for Centiloid measurement error.^19–22^ In the homogeneous scenarios, individuals shared the rate parameter (*k*), sampled for each simulated dataset from a Uniform distribution. In the heterogeneous scenarios, individuals’ rate parameters could vary randomly around the simulated dataset’s mean rate.

Simulated Centiloid measurements under the logistic accumulation scenarios followed

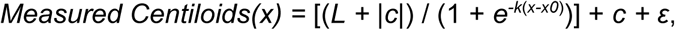

where *L* is the function maximum (ceiling), *c* is an offset that sets the function minimum (≤ 0), and the rest of the terms are defined as specified for the exponential law. In the homogeneous scenarios, individuals shared the rate (*k*) and ceiling (*L*) parameters, which were sampled for each simulated dataset from Uniform distributions. In the heterogeneous scenarios, individuals’ rate and ceiling parameters could vary randomly around these parameters.

For both the exponential and logistic heterogeneous cases, each individual’s value was sampled from a Normal distribution with a standard deviation of 10% the simulated dataset’s rate and, if applicable, ceiling parameters. We also included offsets to produce minimum “true” Centiloid values around −39, the lowest recorded Centiloid measurement in our ADNI sample.

In secondary analyses, we generated linear amyloid accumulation trajectories. Simulation parameters, distributions, and calculations appear in **Supplementary Table 2** and **Supplementary Methods.**

### SILA Fits to Simulated Data

We fit SILA to each simulated dataset and extracted its estimates of AAPO and Centiloid values. For the exponential amyloid accumulation trajectories, we dropped simulated Centiloid values greater than 350 prior to fitting SILA, similar to the maximum Centiloid value in our ADNI sample (346). We plotted SILA’s predictions for each dataset, stratifying by simulation scenario, to gauge its ability to capture the shapes of the “true” population average trajectories. All analyses were conducted using R version 4.5.3.

### Ethics Statement

While this project was approved by the Brown Institutional Review Board, the current analyses used de-identified data from ADNI and are not considered human subjects research. ADNI participants provided informed consent at the time of enrollment, and IRB approval was obtained at each site.

## Results

### SILA in ADNI

Characteristics of the ADNI sample appear in **Table 1**. Centiloid measurements across available PET scans ranged from −39 to 346.

When we plotted SILA’s predicted Centiloids against estimated AAPO, we observed a gradual incline prior to amyloid positivity and a steep incline thereafter, with neither an apparent amyloid ceiling nor a slowing of amyloid accumulation (**Figure 1**). The maximum time elapsed from estimated AAPO was approximately 32 years, though 95% of estimated times were below 16.7 years.

**Figure 1.**
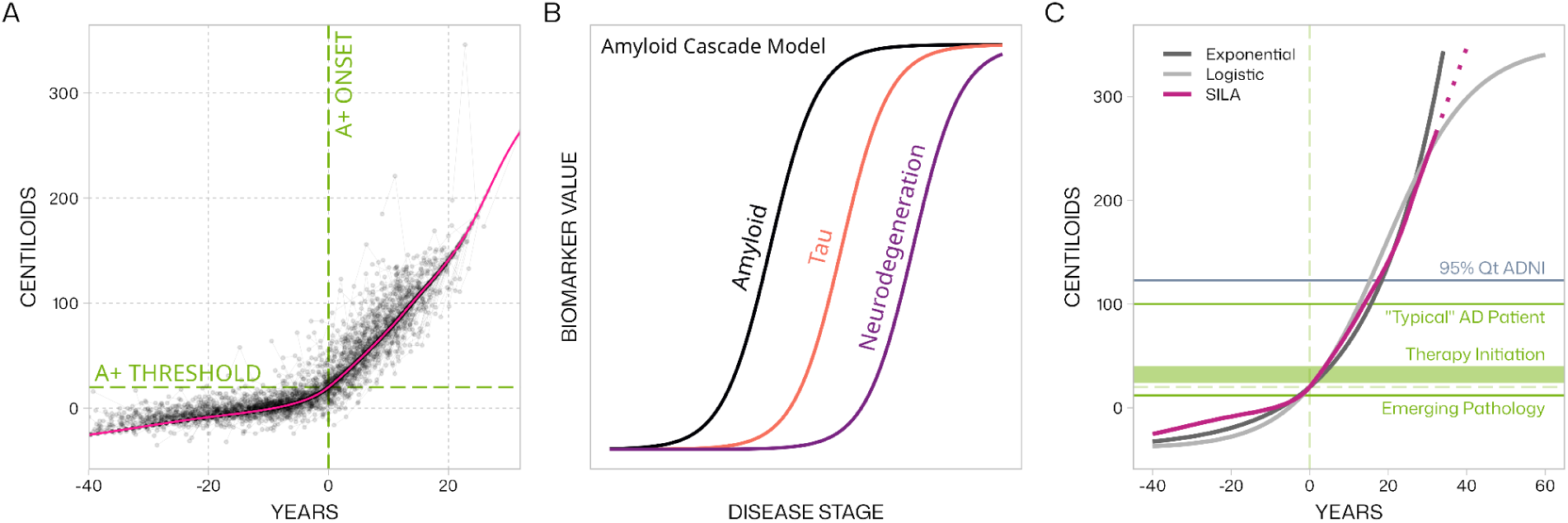
SILA predictions based on a fit in empirical data from ADNI (A) alongside a hypothetical sigmoid model of biomarker cascade (B) and a comparison of SILA against exponential and logistic functions (C). In Panel A, the pink line represents SILA’s predicted Centiloids relative to its prediction of the individual’s age at amyloid positivity onset. Gray dots depict empirical Centiloid measurements from ADNI (3,539 PET scans; 1,141 subjects), with gray lines joining observations from the same subject. The dashed horizontal green line depicts the amyloid positivity onset threshold (20 Centiloids), while the dashed vertical green line depicts the time of amyloid positivity onset. In Panel B, we depict a hypothetical amyloid cascade model in which biomarkers accumulate sequentially, each taking on a sigmoid (logistic) shape in the evolution of their magnitudes, similar to models that have been proposed.^8,10^ In Panel C, we show SILA’s predicted accumulation trajectory alongside exponential and logistic functions of similar magnitude. The dotted pink line depicts a linear extrapolation of SILA’s prediction. Dashed vertical and horizontal green lines follow from Panel A. We augment the plot to show the amyloid load of a “typical” AD patient (100 Centiloids),^28^ a range of thresholds considered adequate for initiating anti-amyloid therapy,^29^ and the point at which emerging amyloid pathology first becomes apparent.^29,30^ AD, Alzheimer’s disease; ADNI, Alzheimer’s Disease Neuroimaging Initiative; SILA, sampled iterative local approximation.

### Simulated SILA Fits

Across all simulation scenarios, SILA captured the qualitative shape of the “true” amyloid accumulation trajectory. That is, when no ceiling was present SILA did not detect one, and when a ceiling was present SILA detected it (**Figure 2**). The impact of inter-individual heterogeneity was more apparent for the logistic laws, where SILA’s Centiloid predictions indicated more gradual changes in the average amyloid trajectory compared to the homogeneous scenarios but still detected the presence of the ceiling. Secondary analyses using linear amyloid accumulation trajectories (ceilingless) did not change inference (**Supplementary Figures 6–9**).

**Figure 2.**
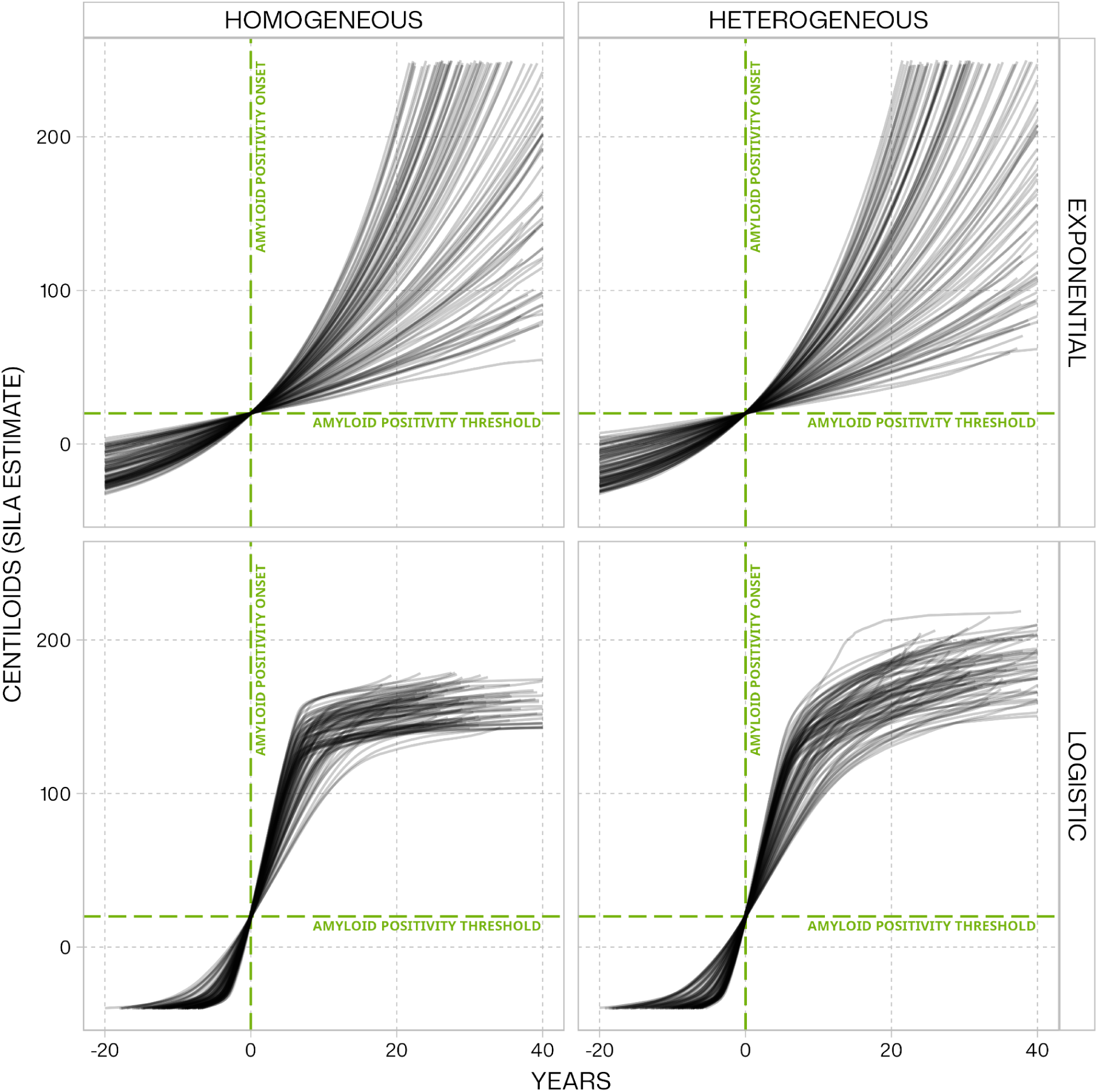
Sampled iterative local approximation (SILA) fits for the exponential and logistic simulation scenarios. All panels depict SILA-estimated Centiloids relative to the SILA-estimated time of amyloid positivity onset under the homogeneous and heterogeneous exponential and logistic amyloid accumulation trajectories. Exponential trajectories varied in the rate of amyloid accumulation, while logistic trajectories varied in both the rate and ceiling of amyloid accumulation. In the homogeneous scenarios, all individuals in a simulation were assigned the same accumulation parameters, whereas the heterogeneous case incorporated inter-individual variation in these parameters. For the sake of differentiating SILA’s predictions in each simulated dataset, the *y*-axis is truncated to suppress predictions below −20 and above 200 Centiloids, and the *x*-axis is truncated to capture the period from 20 years prior to 40 years after estimated amyloid positivity onset. The simulated exponential trajectories in particular could produce extreme “true” Centiloid values. Each panel represents SILA estimates from 100 simulated datasets.

## Discussion

When fit to simulated datasets SILA reliably detected a ceiling in amyloid accumulation when one was present and recovered the correct qualitative shapes of trajectories even when the method’s homogeneity assumptions were violated. SILA did not detect an amyloid ceiling in the ADNI sample, consistent with previous fits in this cohort.^1^

Betthauser et al. fit SILA and other estimators on data from ADNI, the Baltimore Longitudinal Study of Aging (BLSA), and the Wisconsin Registry for Alzheimer’s Prevention (WRAP), finding that SILA appeared not to detect a physiologic ceiling when plotting Centiloids against estimated time since AAPO.^1^ When change in amyloid was plotted against within-person global means, amyloid accumulation appeared to increase with time without hitting a ceiling in ADNI. While accumulation rates did appear to level off in BLSA and WRAP, there was substantial imprecision due to sparse sampling at high amyloid levels. Sparsity may have also amplified partial volume effects related to accelerating brain atrophy, which can yield underestimation of amyloid burden and could produce an apparent ceiling.^6^ The SILA-estimated trajectory in ADNI is likely more robust due to a larger sample size overall; it may also more accurately reflect amyloid trajectories across the agespan and AD continuum because ADNI participants are older on average, with higher rates of AD dementia compared to BLSA and WRAP. Together, these factors suggest that if a physiologic ceiling were present it should be detectable in ADNI.

By simulating datasets similar to ADNI and then generating amyloid trajectories under these constraints, we produced realistic datasets under a range of true amyloid accumulation trajectories. We found that SILA reliably captures the qualitative shape of the average amyloid trajectory across different hypothetical accumulation scenarios, and that SILA-estimated mean trajectories are likely robust to inter-individual variability in amyloid accumulation rates.

Having determined that SILA should reliably identify an amyloid ceiling if present, it is notable that our results in the empirical data corroborate the authors’ original findings in a larger ADNI sample with longer follow-up than was available in 2022. Within the 20 years after estimated amyloid onset (where observed data were not overly sparse), SILA-estimated amyloid accumulation showed no apparent signs of leveling off (**Figure 1**). Although a theoretical amyloid ceiling may exist, severe cognitive impairment that precludes PET measurement and elevated mortality rates after AD dementia onset may render a ceiling unobservable and perhaps less clinically informative during the typical course of disease.^23^ Illustrative models of amyloid accumulation that focus on physiologic ceilings^8,9^ are useful thinking tools but may fail to capture temporal dynamics important to informing prevention and treatment interventions during a patient’s disease course.^24^

The simulations we used to support our conclusions incorporated a range of amyloid accumulation trajectories designed to reflect uncertainties regarding trajectory shapes and to examine SILA’s robustness to violations of one of its key assumptions. While not exhaustive, our simulations indicate that SILA’s predictions likely provide qualitatively accurate inferences regarding the presence or absence of a physiologic ceiling, the possible exception being in the case of a population with slow, heterogeneous accumulation rates (**Supplementary Figure 7**). Individuals pre-AAPO generally appear to fit this description, and therefore, using SILA to estimate AAPO among all individuals in the ADNI to inform the corresponding simulation parameters could be viewed as a limitation. However, we intended our simulations were to represent the whole ADNI sample, not just those with longitudinal PET measurements. We also did not incorporate variation due to other factors, such as APOE-ε4 allele carriage, that could affect the timing or course of amyloid accumulation.^1,25–27^ If the presence of a physiologic ceiling is found to vary by prognostic or etiologic factors, SILA analyses could be stratified by a given factor of interest. We did observe some cases, primarily in the heterogeneous logistic scenarios, in which the presence of a physiologic ceiling is obscured somewhat by inter-individual variation (**Figure 3**). In this case, the population average is a less informative prediction for the individual’s amyloid trajectory, with some individuals hitting a ceiling very early or having a lower maximum (*L*) and vice versa. One alternative explanation for the apparent absence of a physiologic ceiling in our empirical analysis is that individuals who reach this ceiling become too ill to undergo follow-up PET scans, but prior studies claiming evidence for a ceiling suffer from this same limitation.

**Figure 3.**
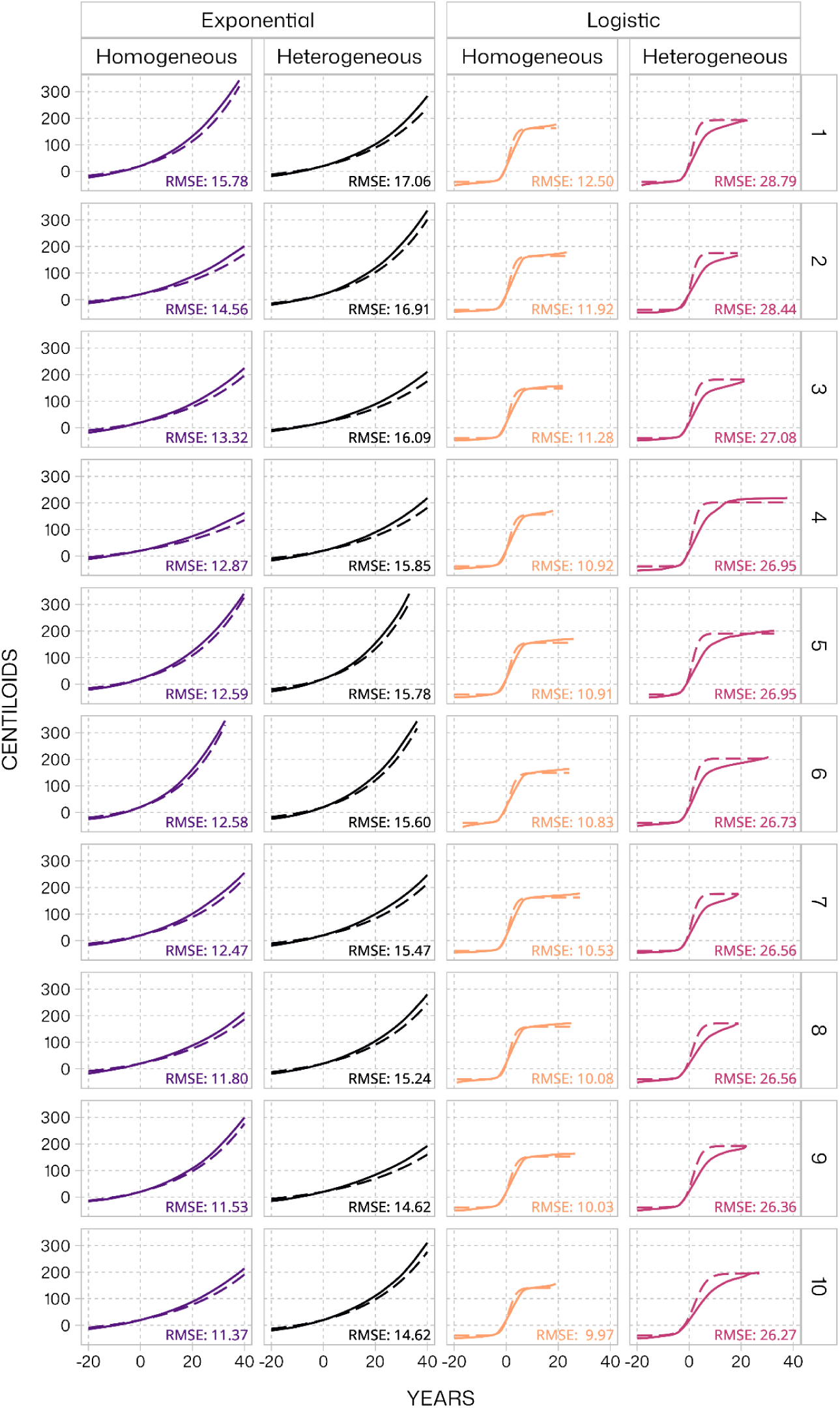
The ten SILA predictions (solid lines) with the highest RMSEs (i.e., worst fits) for the population average amyloid accumulation trajectories (dashed lines) in the exponential and logistic amyloid accumulation scenarios. RMSE calculated at each timepoint for which SILA produced a prediction. Amyloid curves are plotted relative to SILA-estimated age at amyloid positivity onset in a given simulated dataset. In the heterogeneous cases, the population mean parameters were used to calculate “true” Centiloids (dashed lines). SILA, sampled iterative local approximation; RMSE, root mean squared error.

In conclusion, we argue that amyloid does not likely reach a physiologic ceiling during the course of AD. When such a ceiling is present in simulated data SILA captures it. When fit to ADNI, the most comprehensive longitudinal amyloid PET data available, SILA predicts that amyloid accumulation continues unabated for at least 20 years after amyloid positivity onset. Future causal and temporal models of AD biomarker evolution should not necessarily assume a sequential cascade in which all biomarkers evolve in a sigmoid shape. Detailed longitudinal data on multiple biomarkers are needed to inform more robust qualitative and quantitative models.

## Prior Presentation

Preliminary results from this work were presented at the Alzheimer’s Association International Conference 2025 in Toronto, Ontario, Canada (July 27–31, 2025).

## Data Availability Statement

ADNI data may be requested via https://adni.loni.usc.edu. Our code and simulated datasets are available at https://github.com/jrgant/paper-sila-ceiling or via Open Science Framework (https://doi.org/10.17605/OSF.IO/B6PJ3).

## Abbreviations

AAPO: age at amyloid positivity onset
AD: Alzheimer’s disease
ADNI: Alzheimer’s Disease Neuroimaging Initiative
BLSA: Baltimore Longitudinal Study of Aging
PET: positron emission tomography
SILA: sampled iterative local approximation
WRAP: Wisconsin Registry for Alzheimer’s Prevention

## Supplement to

**Supplementary Table 1.** Subsets of ADNI and Berkeley scan data used to estimate simulation input parameters for cohort generation.

| Parameter | Subjects (N) | Scans (N) | Description<sup>a</sup> |
| --- | --- | --- | --- |
| Number of scans<br>Age at first scan | 2,162 | 4,561 | All subjects in the Berkeley scan data with at least one PET scan. |
| Time between scans | 1,142 | 2,399 | Subjects in the Berkeley scan data with at least 2 PET scans. Defined only for scans beyond the first. |
| Age at amyloid positivity onset | 2,149 | 3,539<sup>b</sup> | Subjects included in the prior step and with at least 1 PET scans. |
| <sup>a</sup> Berkeley scan data: University of California, Berkeley processing pipeline (download date: 6mm, February 13, 2026). PTDEMOG and DXSUM (download version: February 13, 2026)<br><sup>b</sup> Omits 1 individual with missing Centiloid measurement at baseline and only one subsequent scan with non-missing Centiloids<br>ADNI, Alzheimer's Disease Neuroimaging Initiative; PET, positron emission tomography |  |  |  |

**Supplementary Table 2.**
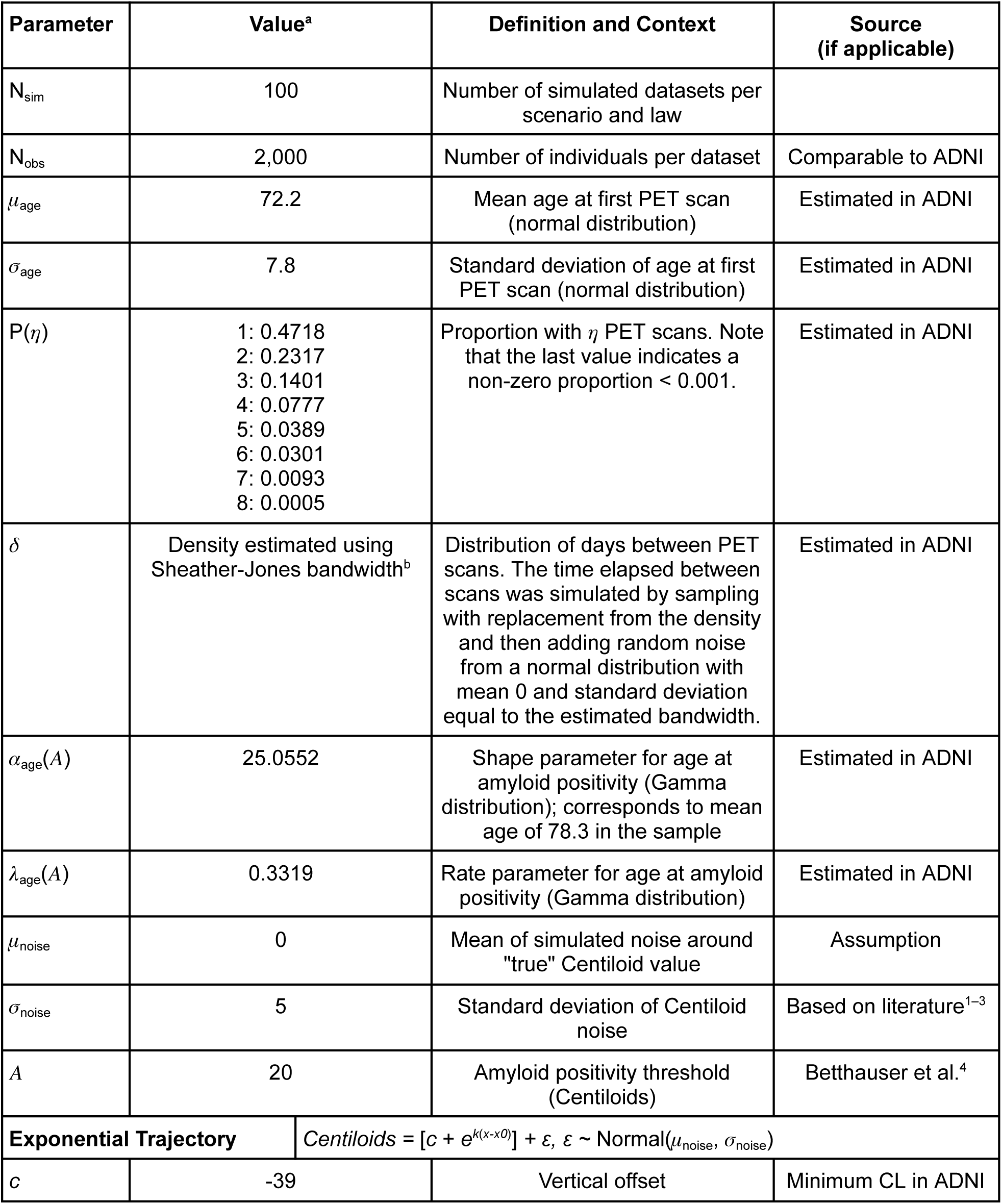
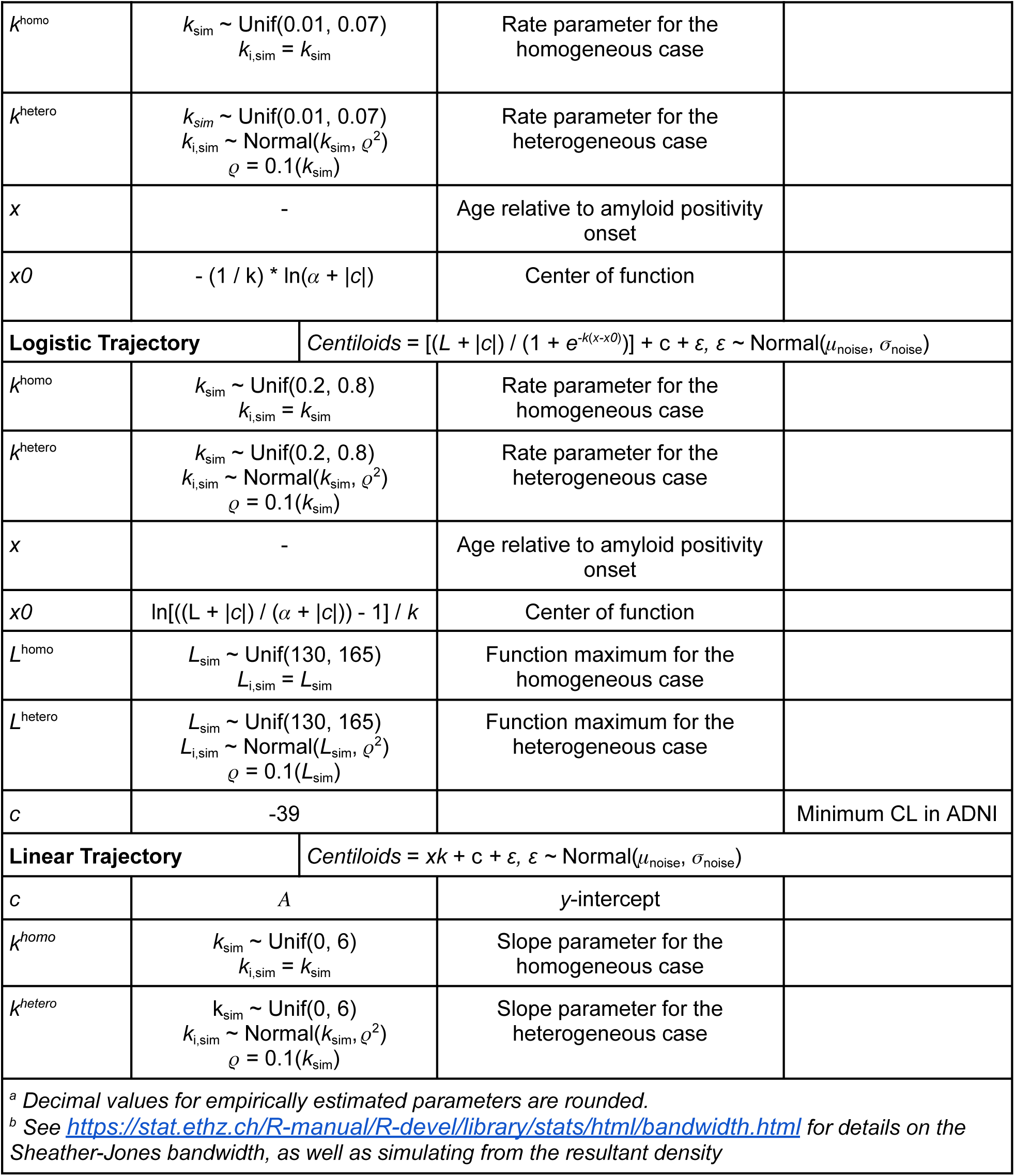
Simulation parameters. Input parameters used to simulate ADNI-like cohorts of individuals and amyloid dynamics according to hypothetical amyloid accumulation trajectories. ADNI, Alzheimer’s Disease Neuroimaging Initiative; CL, Centiloids.

### Supplementary Methods

Here we present the general steps taken during the simulation of a given dataset. Parameter symbols are defined in Supplementary Table 2.

1. Initialize the dataset as follows:

a. Sample with replacement from P(𝜂) to determine how many PET scans each simulated individual receives.
b. Sample from a Gamma distribution with shape 𝛼_age_(𝛢) and rate 𝜆_age_(𝛢), to generate the age at amyloid positivity onset.
c. Sample from a Normal distribution with mean 𝜇_age_ and standard deviation 𝜎_age_ to generate the age at first scan.
2. Convert the dataset to long format so that it contains one row per simulated individual and PET scan.
3. For each scan after the first, sample from 𝛿 to determine the number of days elapsed since the prior scan.
4. Calculate time zero as the age at first scan minus the age at amyloid positivity onset.
5. Calculate the time of each scan relative to disease onset to get *x*.
6. Use the desired amyloid trajectory generation function to get the “true” Centiloids as a function of *x*.
7. Draw from a Normal distribution with mean 0 and standard deviation 𝜎_noise_ and add the result to the “true” Centiloids to get the simulated Centiloid measurement.

**Supplementary Figure 1.**
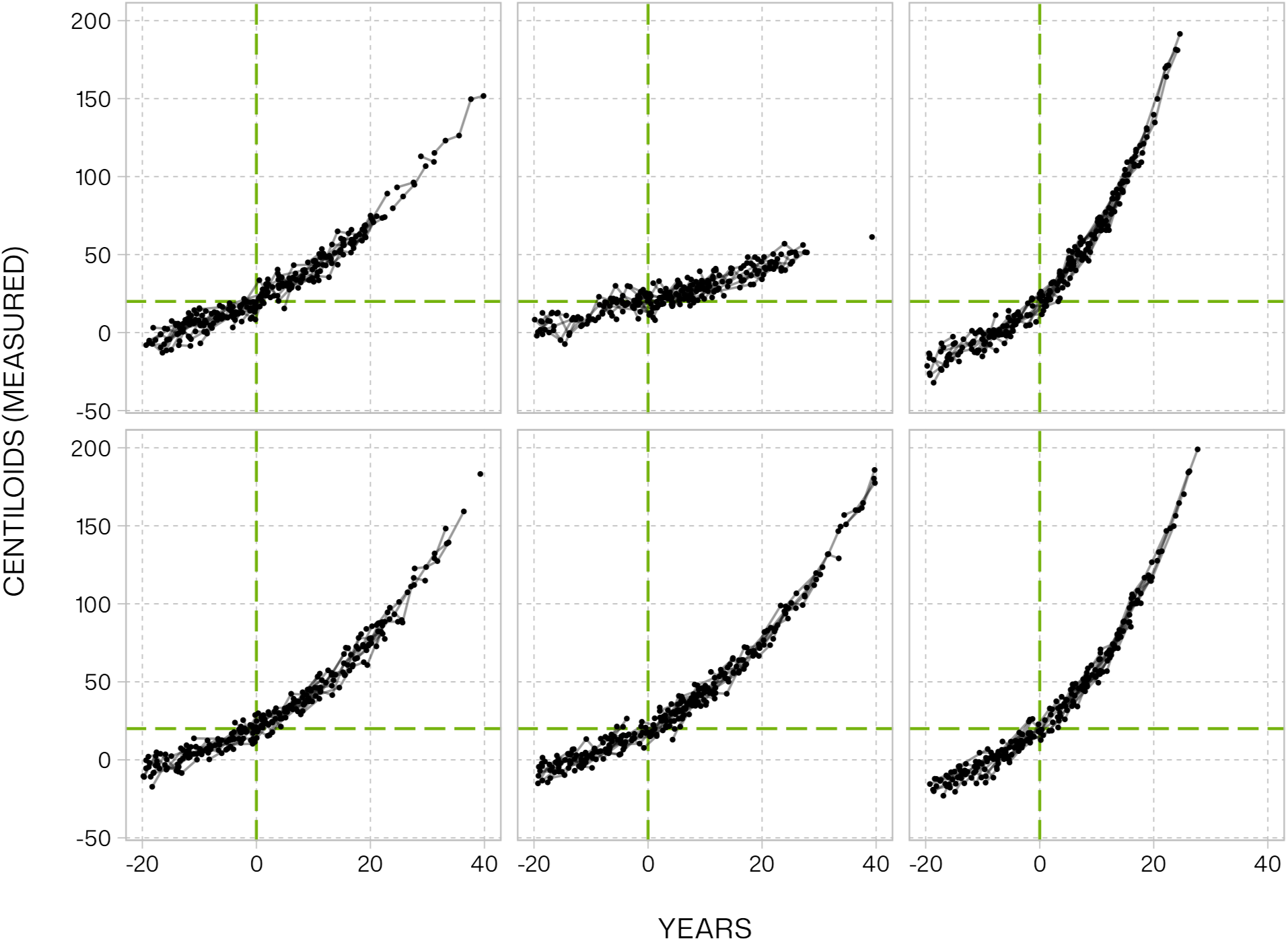
Simulated amyloid observations under the homogeneous exponential trajectories. A random draw of Centiloid measurements from 6 simulated datasets and 100 subjects per dataset. Lines join Centiloid measurements within subjects. The *y*-axis is truncated to a maximum of 200 Centiloids and the *x*-axis to a minimum of −20 years and a maximum of 40 years relative to amyloid positivity onset, in order to better visualize individual observations. The green vertical dashed line indicates the time of amyloid positivity onset and the green horizontal dashed line the amyloid positivity threshold.

**Supplementary Figure 2.**
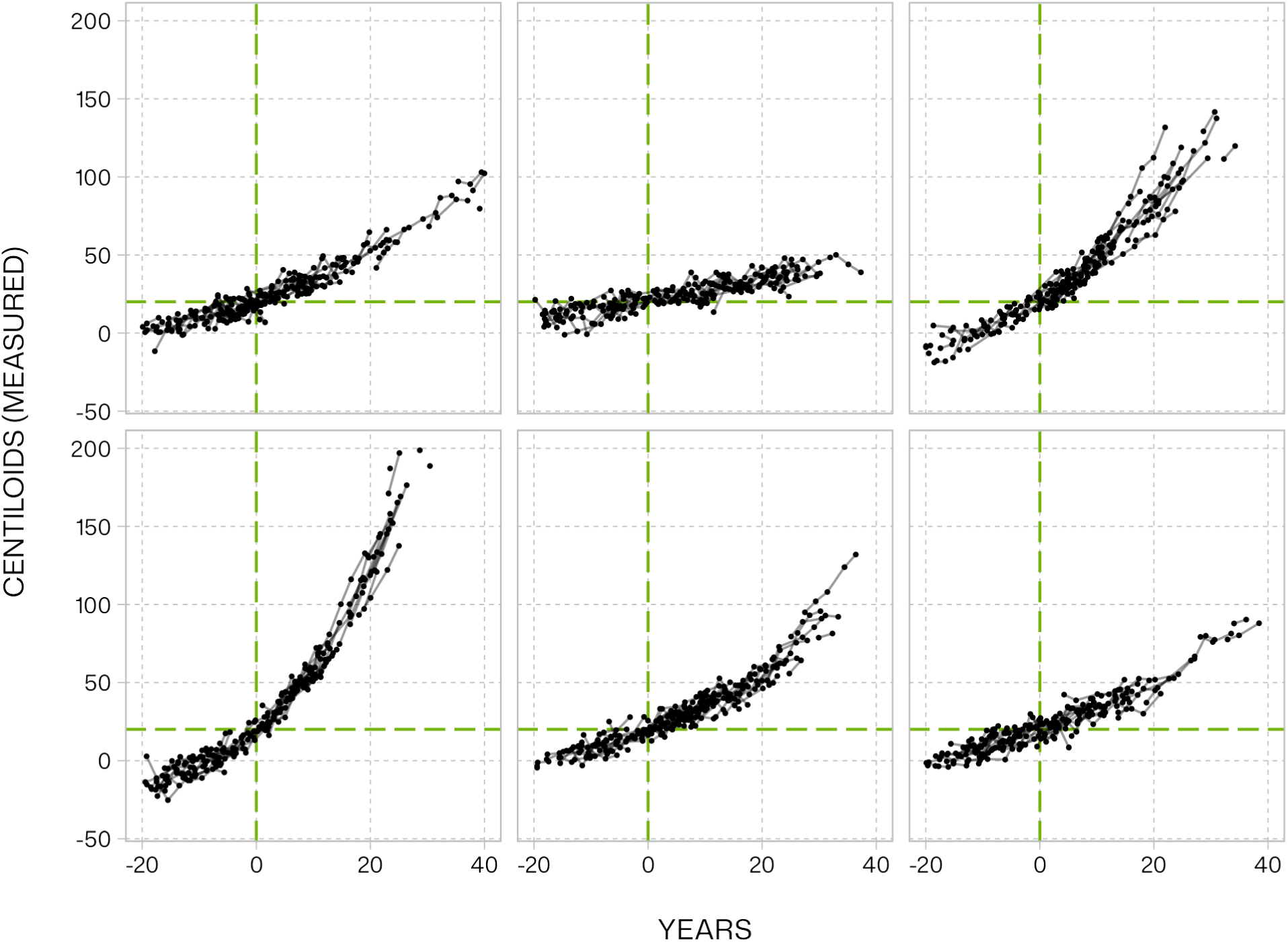
Simulated amyloid observations under the heterogeneous exponential trajectories. A random draw of Centiloid measurements from 6 simulated datasets and 100 subjects per dataset. Lines join Centiloid measurements within subjects. The *y*-axis is truncated to a maximum of 200 Centiloids and the *x*-axis to a minimum of −20 years and a maximum of 40 years relative to amyloid positivity onset, in order to better visualize individual observations. The green vertical dashed line indicates the time of amyloid positivity onset and the green horizontal dashed line the amyloid positivity threshold.

**Supplementary Figure 3.**
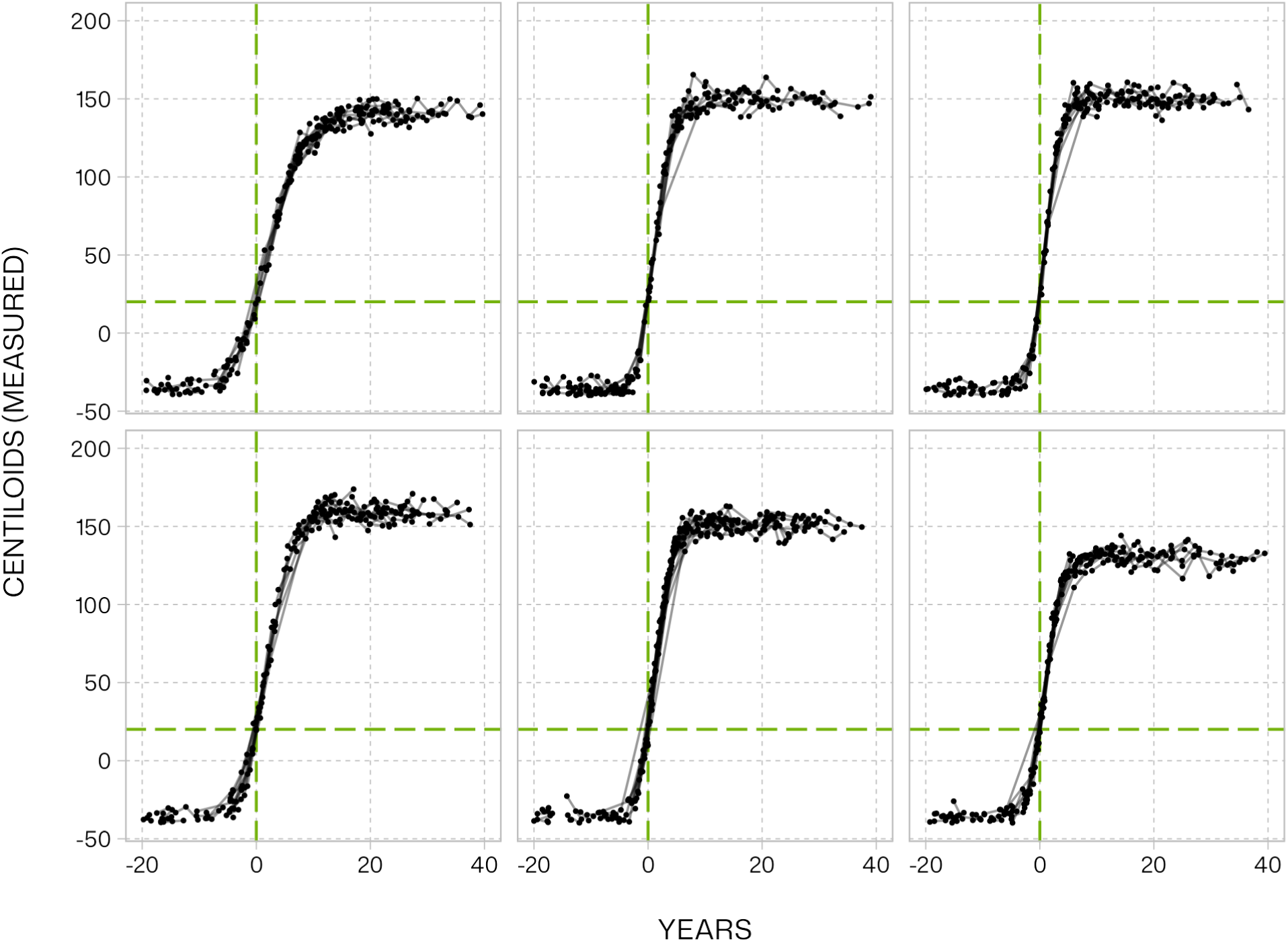
Simulated amyloid observations under the homogeneous logistic trajectories. A random draw of Centiloid measurements from 6 simulated datasets and 100 subjects per dataset. Lines join Centiloid measurements within subjects. The *y*-axis is truncated to a maximum of 200 Centiloids and the *x*-axis to a minimum of −20 years and a maximum of 40 years relative to amyloid positivity onset, in order to better visualize individual observations. The green vertical dashed line indicates the time of amyloid positivity onset and the green horizontal dashed line the amyloid positivity threshold.

**Supplementary Figure 4.**
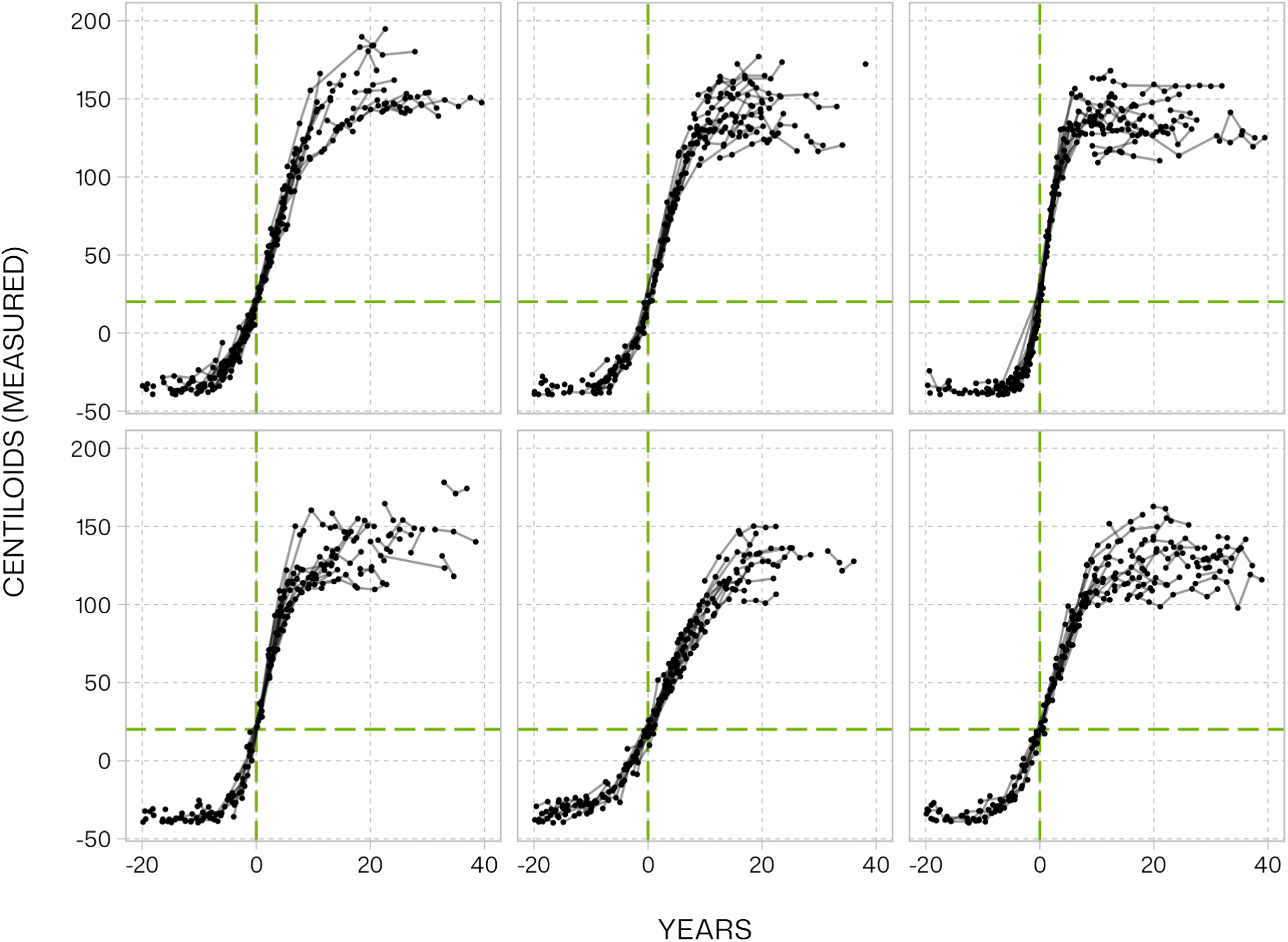
Simulated amyloid observations under the heterogeneous logistic trajectories. A random draw of Centiloid measurements from 6 simulated datasets and 100 subjects per dataset. Lines join Centiloid measurements within subjects. The *y*-axis is truncated to a maximum of 200 Centiloids and the *x*-axis to a minimum of −20 years and a maximum of 40 years relative to amyloid positivity onset, in order to better visualize individual observations. The green vertical dashed line indicates the time of amyloid positivity onset and the green horizontal dashed line the amyloid positivity threshold.

**Supplementary Figure 5.**
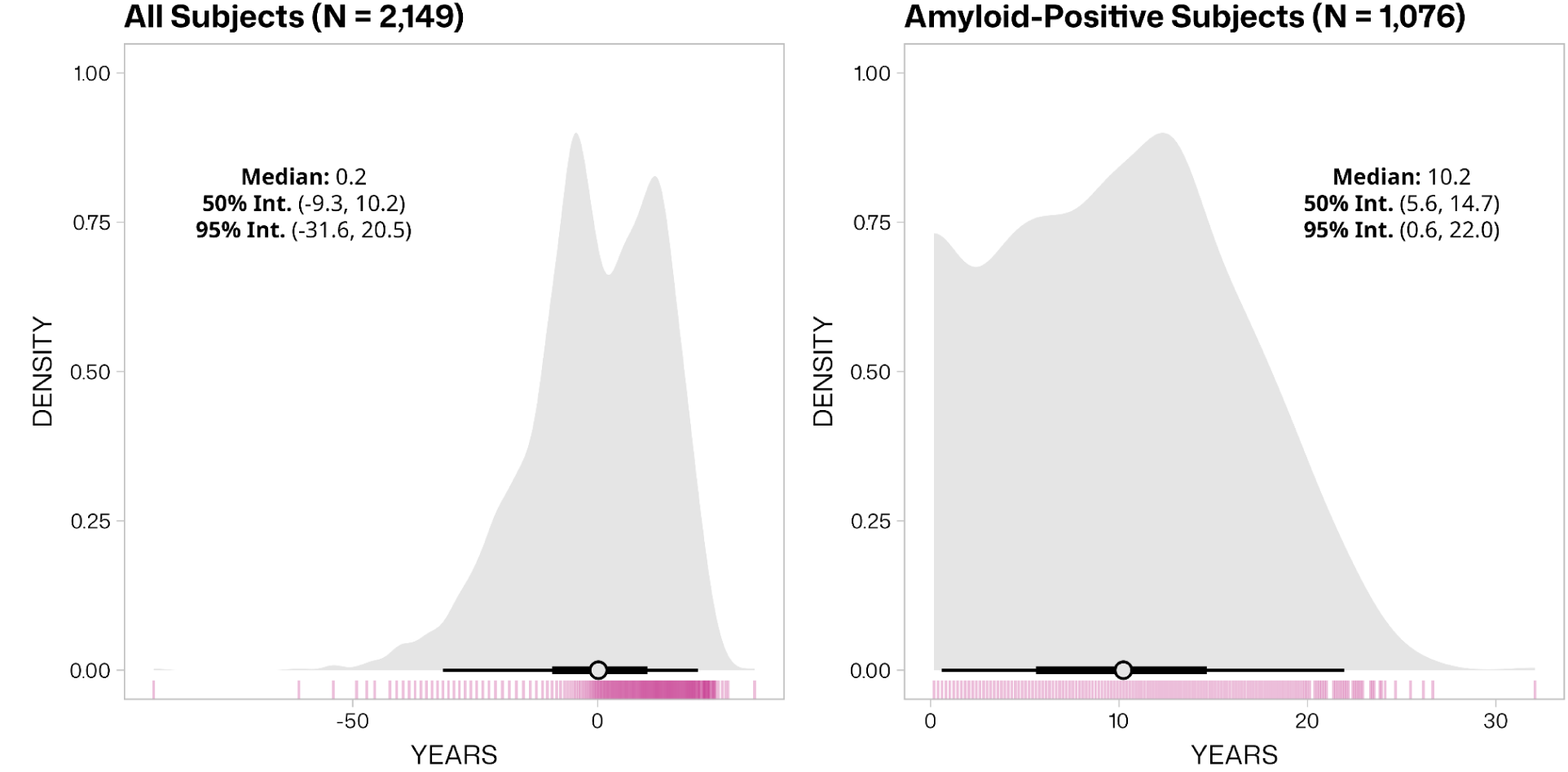
Distribution of years since amyloid positivity onset at individuals’ final PET scan, as estimated by SILA in the ADNI sample, among all subjects and among those with at least one SILA estimate placing them post amyloid positivity onset. Slab intervals depict the middle 50% and 95% of estimates within each panel. PET, positron emission tomography; SILA, sampled iterative local approximation; ADNI, Alzheimer’s Disease Neuroimaging Initiative.

**Supplementary Figure 6.**
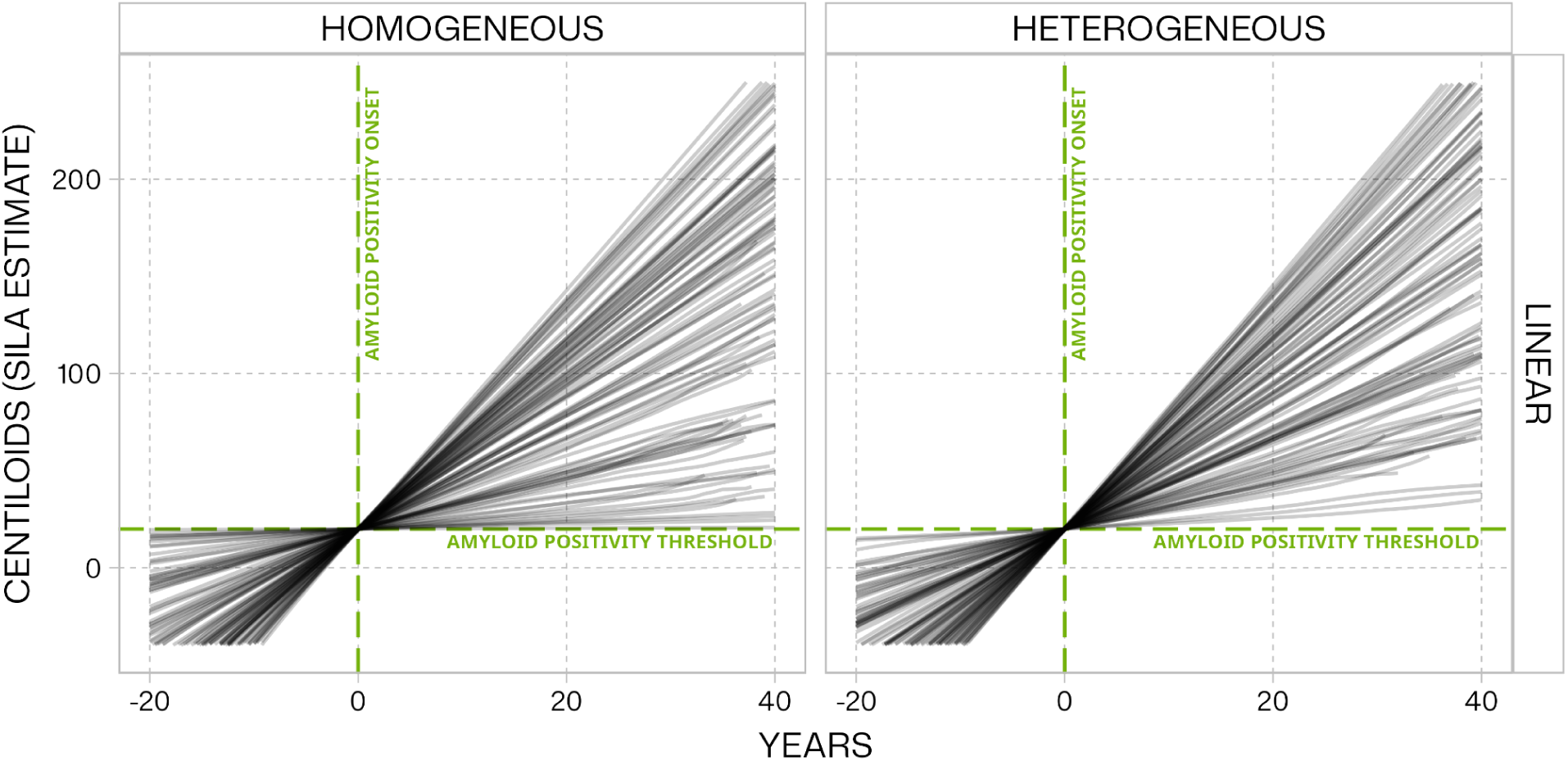
Sampled iterative local approximation (SILA) for the linear simulation scenarios. All panels depict SILA-estimated Centiloids relative to the SILA-estimated time of amyloid positivity onset (20 Centiloids) under the homogeneous and heterogeneous exponential and logistic amyloid accumulation trajectories. Linear trajectories varied in the rate (slope) of amyloid accumulation In the homogeneous case, all individuals in a simulation were assigned the same accumulation parameters whereas the heterogeneous case incorporated inter-individual variation in these parameters. For the sake of differentiating SILA’s predictions in each simulated dataset, the *y*-axis is truncated to suppress predictions below −20 and above 200 Centiloids, and the *x*-axis is truncated to capture the period from 20 years prior to 40 years after amyloid positivity onset. Each panel represents SILA estimates from 100 simulated datasets.

**Supplementary Figure 7.**
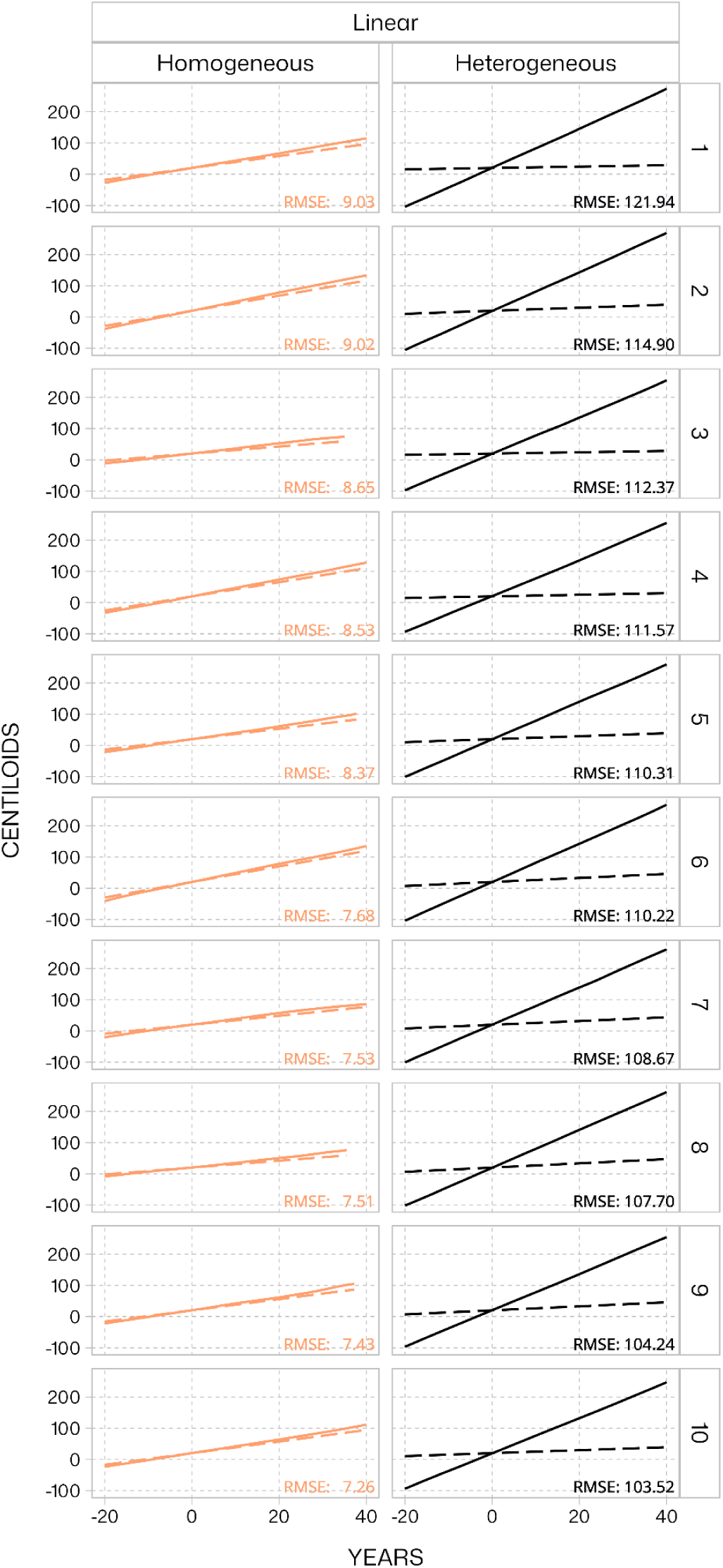
Ten SILA predictions (solid lines) with the highest RMSE for the linear amyloid accumulation scenario. RMSE calculated at each timepoint for which SILA produced a prediction. Amyloid curves are plotted relative to SILA-estimated age at amyloid positivity onset. In the heterogeneous cases, the population mean parameters were used to calculate “true” Centiloids (dashed lines). SILA, sampled iterative local approximation; RMSE, root mean squared error.

**Supplementary Figure 8.**
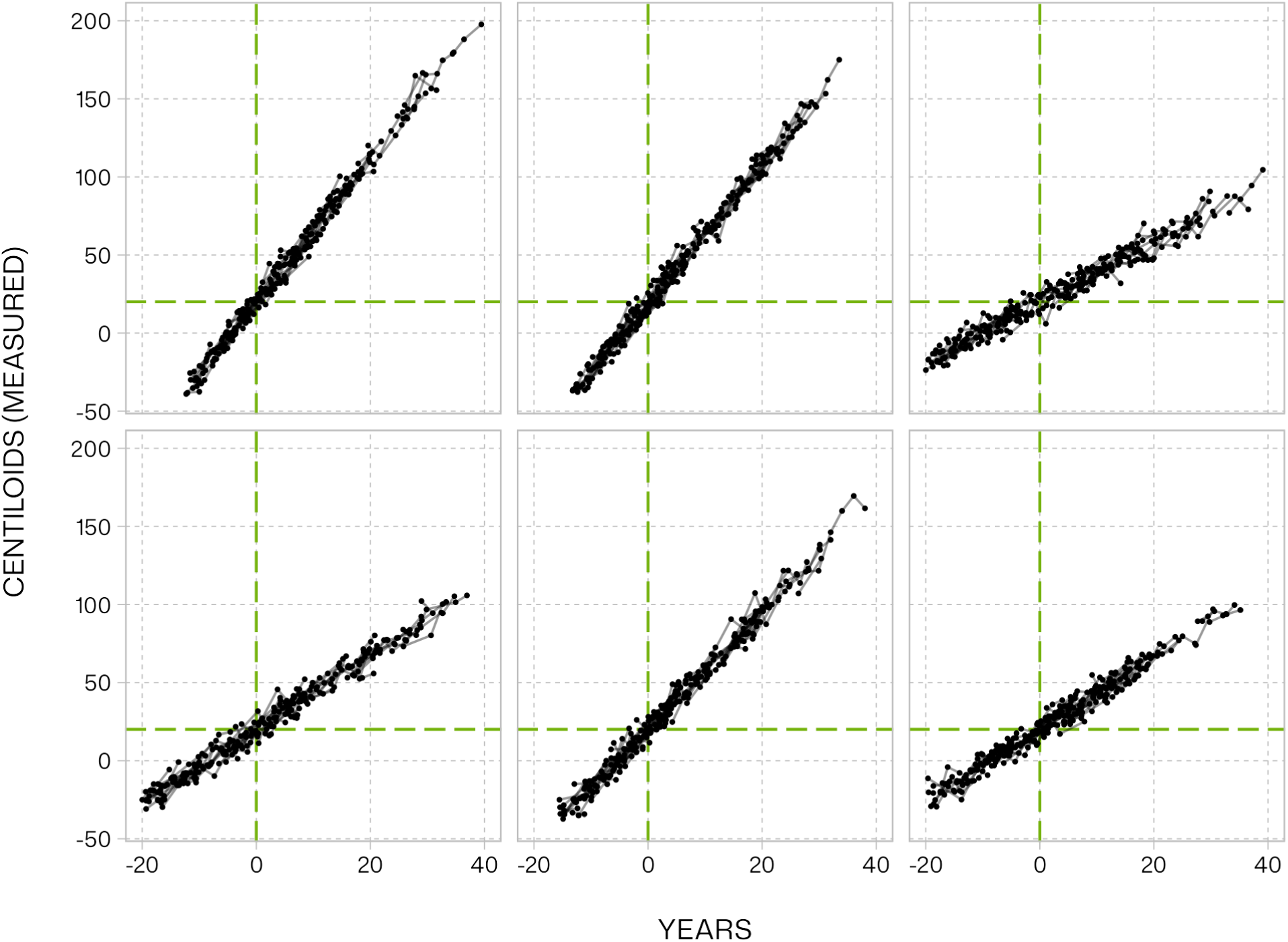
Simulated amyloid observations under the homogeneous linear trajectories. A random draw of Centiloid measurements from 6 simulated datasets and 100 subjects per dataset. Lines join Centiloid measurements within subjects. The *y*-axis is truncated to a maximum of 200 Centiloids and the *x*-axis to a minimum of −20 years and a maximum of 40 years relative to amyloid positivity onset, in order to better visualize individual observations. The green vertical dashed line indicates the time of amyloid positivity onset and the green horizontal dashed line the amyloid positivity threshold.

**Supplementary Figure 9.**
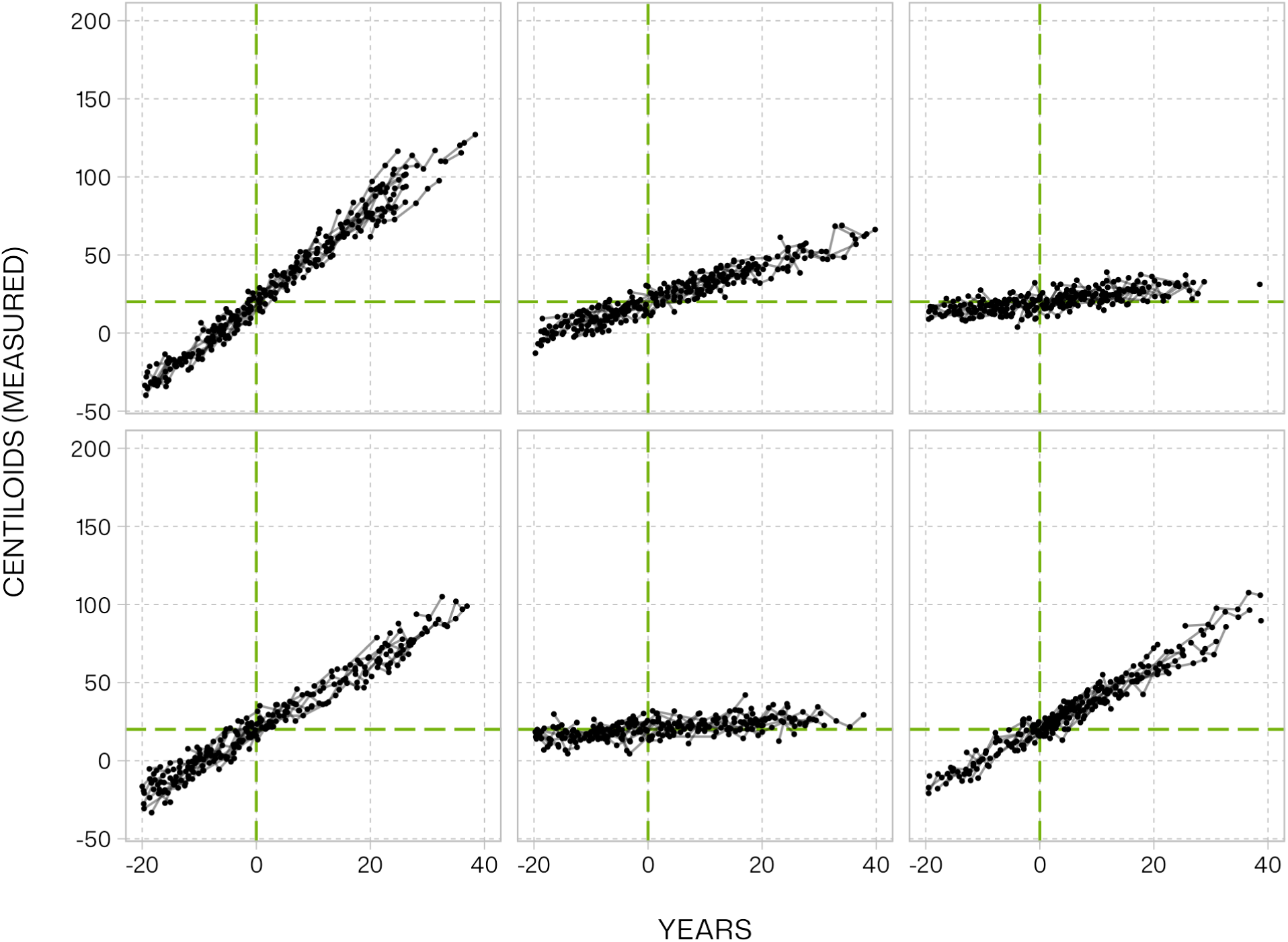
Simulated amyloid observations under the heterogeneous linear trajectories. A random draw of Centiloid measurements from 6 simulated datasets and 100 subjects per dataset. Lines join Centiloid measurements within subjects. The *y*-axis is truncated to a maximum of 200 Centiloids and the *x*-axis to a minimum of −20 years and a maximum of 40 years relative to amyloid positivity onset, in order to better visualize individual observations. The green vertical dashed line indicates the time of amyloid positivity onset and the green horizontal dashed line the amyloid positivity threshold.

## References

1. Betthauser TJ, Bilgel M, Koscik RL, et al. Multi-method investigation of factors influencing amyloid onset and impairment in three cohorts. Brain. 2022;146(2):e11–e11. doi:10.1093/brain/awac213

2. Jedynak BM, Lang A, Liu B, et al. A computational neurodegenerative disease progression score: Method and results with the Alzheimer’s disease neuroimaging initiative cohort. NeuroImage. 2012;63(3):1478–1486. doi:10.1016/j.neuroimage.2012.07.059

3. Therneau TM, Knopman DS, Lowe VJ, et al. Relationships between β-amyloid and tau in an elderly population: An accelerated failure time model. NeuroImage. 2021;242:118440. doi:10.1016/j.neuroimage.2021.118440

4. Schindler SE, Li Y, Buckles VD, et al. Predicting Symptom Onset in Sporadic Alzheimer Disease With Amyloid PET. Neurology. 2021;97(18). doi:10.1212/WNL.0000000000012775

5. Insel PS, Donohue MC, Berron D, Hansson O, Mattsson-Carlgren N. Time between milestone events in the Alzheimer’s disease amyloid cascade. NeuroImage. 2021;227:117676. doi:10.1016/j.neuroimage.2020.117676

6. Villemagne VL, Burnham S, Bourgeat P, et al. Amyloid β deposition, neurodegeneration, and cognitive decline in sporadic Alzheimer’s disease: a prospective cohort study. Lancet Neurol. 2013;12(4):357–367. doi:10.1016/S1474-4422(13)70044-9

7. Staffaroni AM, Quintana M, Wendelberger B, et al. Temporal order of clinical and biomarker changes in familial frontotemporal dementia. Nat Med. 2022;28(10):2194–2206. doi:10.1038/s41591-022-01942-9

8. Jack CR, Knopman DS, Jagust WJ, et al. Hypothetical model of dynamic biomarkers of the Alzheimer’s pathological cascade. Lancet Neurol. 2010;9(1):119–128. doi:10.1016/s1474-4422(09)70299-6

9. Jack CR, Knopman DS, Jagust WJ, et al. Tracking pathophysiological processes in Alzheimer’s disease: an updated hypothetical model of dynamic biomarkers. Lancet Neurol. 2013;12(2):207–216. doi:10.1016/s1474-4422(12)70291-0

10. Jack CR, Wiste HJ, Lesnick TG, et al. Brain β-amyloid load approaches a plateau. Neurology. 2013;80(10):890–896. doi:10.1212/wnl.0b013e3182840bbe

11. Ingelsson M, Fukumoto H, Newell KL, et al. Early Aβ accumulation and progressive synaptic loss, gliosis, and tangle formation in AD brain. Neurology. 2004;62(6):925–931. doi:10.1212/01.WNL.0000115115.98960.37

12. Haneuse S, Schildcrout J, Crane P, Sonnen J, Breitner J, Larson E. Adjustment for Selection Bias in Observational Studies with Application to the Analysis of Autopsy Data. Neuroepidemiology. 2009;32(3):229–239. doi:10.1159/000197389

13. Caroli A, Frisoni GB. The dynamics of Alzheimer’s disease biomarkers in the Alzheimer’s Disease Neuroimaging Initiative cohort. Neurobiol Aging. 2010;31(8):1263–1274. doi:10.1016/j.neurobiolaging.2010.04.024

14. Jagust WJ, Landau SM, for the Alzheimer’s Disease Neuroimaging Initiative. Temporal Dynamics of β-Amyloid Accumulation in Aging and Alzheimer Disease. Neurology. 2021;96(9). doi:10.1212/WNL.0000000000011524

15. Heston MB, Teague JP, Cody KA, et al. Factors associated with age at tau pathology onset and time from tau onset to dementia in Alzheimer’s disease. Alzheimers Dement. 2025;21(8):e70551. doi:10.1002/alz.70551

16. Brown CA, Cousins KAQ, Korecka M, et al. Temporal Modeling of Amyloid and Tau Trajectories in Alzheimer’s Disease Using PET and Plasma Biomarkers. Ann Neurol. Published online March 15, 2026:ana.78194. doi:10.1002/ana.78194

17. Cody KA, Langhough RE, Zammit MD, et al. Characterizing brain tau and cognitive decline along the amyloid timeline in Alzheimer’s disease. Brain. 2024;147(6):2144–2157. doi:10.1093/brain/awae116

18. Landau S, Lee JY, Taggett J, Jagust W. UC Berkeley Amyloid PET Processing Methods. Published online June 20, 2025.

19. Collij LE, Bollack A, La Joie R, et al. Centiloid recommendations for clinical context-of-use from the AMYPAD consortium. Alzheimer’s Dement. Published online November 2024. doi:10.1002/alz.14336

20. Joshi AD, Pontecorvo MJ, Clark CM, et al. Performance Characteristics of Amyloid PET with Florbetapir F 18 in Patients with Alzheimer’s Disease and Cognitively Normal Subjects. J Nucl Med. 2012;53(3):378–384. doi:10.2967/jnumed.111.090340

21. Roé-Vellvé N, Farrar G, Collij L, et al. Impact of error propagation in the development of the centiloid conversion equation. Annu Congr Eur Assoc Nucl Med. 2022;49(Suppl 1):S548.

22. Schmidt ME, Chiao P, Klein G, et al. The influence of biological and technical factors on quantitative analysis of amyloid PET: Points to consider and recommendations for controlling variability in longitudinal data. Alzheimer’s Dement. 2014;11(9):1050–1068. doi:10.1016/j.jalz.2014.09.004

23. Crowell V, Reyes A, Zhou SQ, Vassilaki M, Gsteiger S, Gustavsson A. Disease severity and mortality in Alzheimer’s disease: an analysis using the U.S. National Alzheimer’s Coordinating Center Uniform Data Set. BMC Neurol. 2023;23(1):302. doi:10.1186/s12883-023-03353-w

24. Fischer L, Parker D, Maboudian S, et al. Longitudinal biomarker studies in human neuroimaging: capturing biological change of Alzheimer’s pathology. Alzheimers Res Ther. 2025;18(1):13. doi:10.1186/s13195-025-01920-6

25. Lim YY, Mormino EC, For the Alzheimer’s Disease Neuroimaging Initiative, et al. *APOE* genotype and early β-amyloid accumulation in older adults without dementia. Neurology. 2017;89(10):1028–1034. doi:10.1212/WNL.0000000000004336

26. Villemagne VL, Pike KE, Chételat G, et al. Longitudinal assessment of Aβ and cognition in aging and Alzheimer disease. Ann Neurol. 2011;69(1):181–192. doi:10.1002/ana.22248

27. Mishra S, Blazey TM, Holtzman DM, et al. Longitudinal brain imaging in preclinical Alzheimer disease: impact of APOE ε4 genotype. Brain. 2018;141(6):1828–1839. doi:10.1093/brain/awy103

28. Klunk WE, Koeppe RA, Price JC, et al. The Centiloid Project: Standardizing quantitative amyloid plaque estimation by PET. Alzheimer’s Dement. 2014;11(1):1. doi:10.1016/j.jalz.2014.07.003

29. Farrar G, Weber CJ, Rabinovici GD. Expert opinion on Centiloid thresholds suitable for initiating anti-amyloid therapy. Summary of discussion at the 2024 spring Alzheimer’s Association Research Roundtable. J Prev Alzheimers Dis. 2025;12(1):100008. doi:10.1016/j.tjpad.2024.100008

30. Salvadó G, Molinuevo JL, Brugulat-Serrat A, et al. Centiloid cut-off values for optimal agreement between PET and CSF core AD biomarkers. Alzheimers Res Ther. 2019;11(1):27. doi:10.1186/s13195-019-0478-z

## References

1. Collij LE, Bollack A, La Joie R, et al. Centiloid recommendations for clinical context-of-use from the AMYPAD consortium. Alzheimer’s Dement. Published online November 2024. doi:10.1002/alz.14336

2. Joshi AD, Pontecorvo MJ, Clark CM, et al. Performance Characteristics of Amyloid PET with Florbetapir F 18 in Patients with Alzheimer’s Disease and Cognitively Normal Subjects. J Nucl Med. 2012;53(3):378–384. doi:10.2967/jnumed.111.090340

3. Roé-Vellvé N, Farrar G, Collij L, et al. Impact of error propagation in the development of the centiloid conversion equation. Annu Congr Eur Assoc Nucl Med. 2022;49(Suppl 1):S548.

4. Betthauser TJ, Bilgel M, Koscik RL, et al. Multi-method investigation of factors influencing amyloid onset and impairment in three cohorts. Brain. 2022;146(2):e11–e11. doi:10.1093/brain/awac213

5. Kay M. ggdist: Visualizations of Distributions and Uncertainty in the Grammar of Graphics. IEEE Trans Vis Comput Graph. Published online 2023:1-11. doi:10.1109/TVCG.2023.3327195

